# Multi-omic transcriptional, brain, and clinical variations in schizophrenia

**DOI:** 10.1101/2023.05.30.23290738

**Authors:** Long-Biao Cui, Shu-Wan Zhao, Ya-Hong Zhang, Kun Chen, Yu-Fei Fu, Ting Qi, Mengya Wang, Jing-Wen Fan, Yue-Wen Gu, Xiao-Fan Liu, Xiao-Sa Li, Wen-Jun Wu, Di Wu, Hua-Ning Wang, Yong Liu, Hong Yin, Martijn P. van den Heuvel, Yongbin Wei

## Abstract

How genetic risk variants may relate to brain abnormalities is crucial for understanding cross-scale pathophysiological mechanisms underlying schizophrenia. The present study identifies brain structural correlates of variation in gene expression in schizophrenia and its clinical significance. Of 43 patients with schizophrenia, RNA-seq data from blood samples, MRI, and clinical assessments were collected, together with data from 60 healthy controls. Gene expression differentiation between schizophrenia and health controls was assessed and cross-referenced to schizophrenia-related genomic variations (GWAS on 76,755 patients and 243,649 controls and GWAS on 22,778 East Asian patients) and brain gene expressions (samples from 559 patients and 175 individuals). Multivariate correlation analysis was employed to examine associations across gene expression, brain volume, and clinical assessments. Differentially expressed genes in blood samples from patients with schizophrenia were significantly enriched for genes previously reported in genome-wide association studies on schizophrenia (*P* = 0.002, false discovery rate corrected) and were associated with gene expression differentiation in the brain (*P* = 0.016, 5,000 permutations). Transcriptional levels of differentially expressed genes were found to significantly correlate with gray matter volume in the frontal and temporal regions of cognitive brain networks in schizophrenia (*q* < 0.05, false discovery rate corrected). A significant correlation was further observed between gene expression, gray matter volume, and performance in the Wechsler Adult Intelligence Scale test (*P* = 0.031). Our findings suggest that genomic variations in schizophrenia are associated with differentiation in the blood transcriptome, which further plays a role in individual variations in macroscale brain structure and cognition.

## Introduction

Schizophrenia (SCZ) is a psychiatric disorder with a high heritability^1^ and is determined by multiple genetic factors, such as common variants,^2^ rare copy number variants,^3^ and rare coding variants.^4^ These genomic variants are known to influence the spatiotemporally dynamic patterns of gene expression in brain tissue,^5^ as evidenced by differential expression and splicing in the brain transcriptome of SCZ.^6^ An altered transcriptional profile in SCZ implicates changes in biological pathways in the central nervous system,^7^ which further associate to macroscale brain structural and functional abnormalities in the disorder.^8^ Integrating cross-scale biological data between genetics and macroscale brain phenotypes becomes central to our understanding of the multifactorial etiology of SCZ and to developing precise assessment and treatment.^8^

Emerging studies linking genetics to neuroimaging have pinpointed the functional consequence of SCZ risk genes in the brain. Polygenic risk for SCZ has, for example, been observed to relate to individual variations in brain morphology^9^ and the structural and functional connectome organization of the brain.^10,11^ Recent studies examining the relationship between transcriptomics and neuroimaging phenotypes have further indicated that the transcription of SCZ risk genes in the brain display overlapping patterns between disease-related vulnerabilities in brain morphology and connectivity.^12-14^ Such observed associations depend however on group averaged patterns of neuroimaging data and gene expression data,^15,16^ which is not sufficient to describe inter-individual transcriptional variations of patients with SCZ. It remains largely unknown whether, and if so how, individual variations in gene expression correlate to brain structural abnormalities in SCZ.

Examining gene expressions from blood samples provides a feasible way to investigate participant-level transcriptomic-neuroimaging associations in vivo considering the correspondence of gene expressions between blood and brain.^17,18^ Therefore, we collected a cross-scale cohort of blood RNA-seq, MRI, and clinical data to investigate inter-individual transcriptomic variations in relation to brain structural variations in SCZ patients and their clinical significance.

## Materials and methods

### Participants

Multi-omic RNA-seq data, MRI data, and symptom severity ratings of 43 SCZ patients and 60 age-, sex-matched healthy controls (HCs) were collected in the present study (Table 1). Patients were recruited from the Department of Psychiatry, Xijing Hospital, Xi’an, China, and controls were recruited from the local community. Inclusion and exclusion criteria were described in detail in the Supplementary Methods. Individuals with SCZ completed a battery of clinical assessments and cognitive tests, including the Positive and Negative Syndrome Scale (PANSS)^19^ assessing the clinical symptom severity, as well as the Wechsler Adult Intelligence Scale revised in China (WAIS-RC)^20^ assessing general cognitive ability. The WAIS-RC was also performed in HCs. The behavioral assessments, brain scanning, and blood collection were performed on the same day. This study was approved by the Institutional Ethics Committee, First Affiliated Hospital of Fourth Military Medical University and all participants provided written informed consent of participation in this study.

### RNA-seq data

#### Data acquisition

Intravenous blood from the upper forearm was drawn from each participant. Blood sample of 2.5 ml was collected in a BD Vacutainer tube and was immediately frozen at −80°C. RNA sequencing was performed using Illumina Novaseq 6000. Low-quality reads were filtered out by fastp^21^ (version 0.18.0) and filtered reads were mapped to human reference genome hg19 using HISAT2.2.4.^22^ Details in RNA-seq data collection and analysis were described in Supplementary Methods.

#### Differential expression analysis

The count data of 20,313 genes representing the number of sequence reads were used in differential expression analysis. Standardized DESeq2 analysis^23^ with fold-changes shrunk by the apeglm package^24^ (implemented in R) was performed, resulting in normalized counts of 17,999 genes and their transcriptional differentiations between SCZ and HC. Fold-change (FC) was obtained for each gene to describe the transcriptional differentiation and a *P* value adjusted for multiple comparisons using the false discovery rate (FDR) Benjamini–Hochberg procedure was assigned. Age and sex were regressed out as covariates of no interest. Differentially expressed genes (DEGs) were identified according to an FDR adjusted *P* value below 0.05.

### MRI data

#### Data acquisition

T1-weighted MRI, diffusion weighted imaging (DWI), and resting-state functional MRI (rsfMRI) data were collected using a GE Discovery MR750 3.0 T scanner. Scanning parameters are described in^25^ and are tabulated in Supplementary Table 1.

#### Morphometric metrics

T1-weighted MRI data were preprocessed using FreeSurfer (version 6.0) to segment brain tissues and reconstruct the cortical mantle. The reconstructed cortical mantle was parcellated into 114 distinct regions (i.e. 57 regions per hemisphere) according to a subdivision of the Desikan-Killiany (DK) atlas.^26,27^ The regional volume automatically produced in the FreeSurfer pipeline was used in the following analysis. The original DK atlas with 68 cortical regions was used for validation purposes (Supplementary Results).

#### Connectivity metrics

Connectome reconstruction was conducted on DWI data and fMRI data using FSL (v6.0)^28^ and CATO (v3.1.2).^29^ Detailed processing steps of DWI data and fMRI data were described in Supplementary Methods. A 114×114 structural connectivity (SC) matrix that describes the streamline density (i.e., the number of streamlines between two regions divided by region volume) and a 114×114 functional connectivity (FC) matrix that describes the correlation on the extracted BOLD time series between every two regions were separately generated for each participant. Regional connectivity strength was computed as the summed strength of all connections of each region.

### Statistical analysis

We performed gene property analysis using MAGMA^30^ to examine to what extent gene expression differentiation is associated with gene-level statistics reported in previously published SCZ GWAS^2^ (Fig. 1A). Permutation testing was also performed to examine whether the observed DEGs (here in blood samples) showed differentiated expression in brain samples in SCZ,^6^ exceeding null distributions of differentiated brain expressions of sets of random genes (Supplementary Methods).

**Figure 1.**
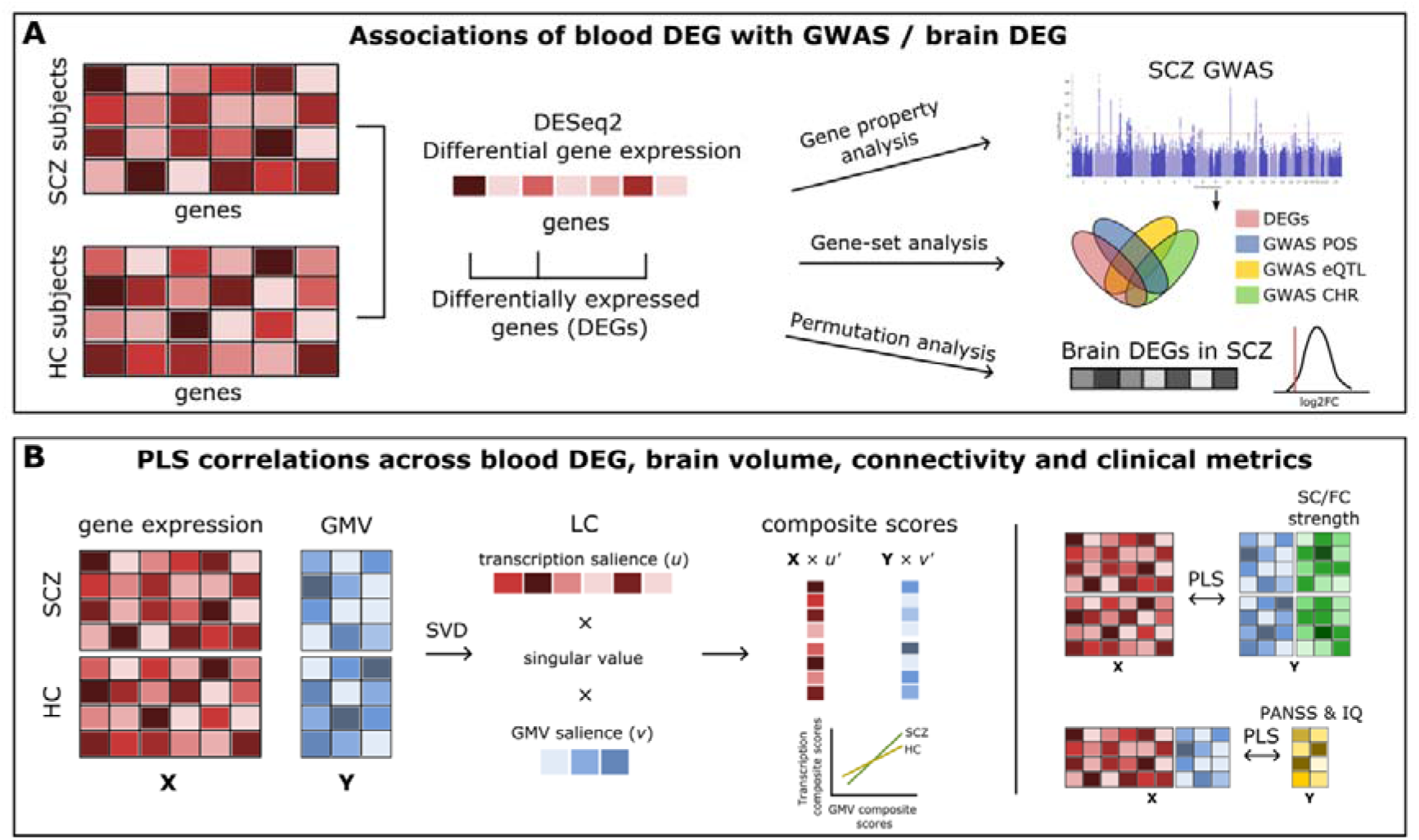
Overview of Methods. **(A)** DESeq2 analysis is performed to examine gene expression differentiation between SCZ and HC. Differentially expressed genes (DEG) are linked to the common genomic variants of SCZ and brain DEG in SCZ. POS: positional mapping; eQTL: expression quantitative trait locus mapping; CHR: chromatin interaction mapping. **(B)** Gene expression level is correlated to brain gray matter volume (GMV) across individuals using PLS analysis. The resulted individual-level composite score is further linked to brain structural and functional connectivity strength and clinical metrics.

Multivariate partial least squares (PLS) correlation analysis was performed to examine associations between gene expressions (i.e., normalized count) of the DEGs and imaging-derived phenotypes (Fig. 1B). PLS has been widely used to seek latent components (LCs) by maximizing the covariance between two data matrices.^31,32^ Age and sex were regressed out as covariates of no interest from the two data matrices using linear regression. Group differences were disregarded by normalizing data within each group separately, such that the observed correlation is not driven by group differences and the common and differing effects of transcription-imaging correlations exist in both SCZ and HC groups.^31,32^

An LC derived from PLS describes a combination of a singular value (indicating the level of the explained variance) and weights (referred to as PLS saliences) for each variable in the input data matrices, e.g., each gene and each brain metric.^31^ Permutation testing (with 5,000 permutations in which subject labels were shuffled) was used to statistically evaluate the observed correlation and bootstrapping (with 5,000 samplings) was further used to estimate whether the resulting PLS saliences of the LCs were robust.^33^ We further performed permutation tests by selecting random genes from the pool of 17,999 genes in each permutation to examine the specificity of SCZ DEGs.^16^ Post hoc analyses on the observed correlation patterns were performed in the context of brain cognitive networks, between-group brain volume differences, and gene expression profiles based on the Allen Human Brain Atlas^34^ (see details in Supplementary Methods). For any LC, the individual-level composite scores^35^ for gene expressions and imaging-derived phenotypes were separately computed to assess the level of covariance captured by the LC. PLS analysis was conducted using myPLS (https://github.com/danizoeller/myPLS).^33^ For validation purposes, the dataset was randomly split into two halves, with PLS analysis separately repeated in each half.

### Data availability

The data supporting the findings of this study are available from the corresponding author upon reasonable request.

## Results

### Differentiated gene expression is associated with schizophrenia-related genomic variations

Gene property analysis showed gene expression differentiation (i.e., log-FC, across 17,999 genes) to be significantly associated with gene-based statistics derived from a recent GWAS on 76,755 SCZ and 243,649 HC^2^ (β = 0.196, *P* = 0.002; FDR corrected; Table 2). A similar result was observed for GWAS on SCZ in the East Asian population^36^ (β = 0.135, *P* = 0.008; FDR corrected; Table 2). These results confirm the genomic risk loci observed for SCZ to be associated with more differentiated gene expression in SCZ compared to HC.^37^

Using GWAS summary statistics from four other common psychiatric disorders (namely bipolar disorder (BD), attention-deficit/hyperactivity disorder (ADHD), autism spectrum disorder (ASD), major depressive disorder (MDD), and insomnia) we examined the level of specificity of this finding. The observed differentiation in gene expressions was found to be significantly associated with genomic variations related to BD^38^ (a condition genetically strongly related to SCZ)^39^ (β = 0.164, *P* = 0.003), but not for ADHD^40^ (β = −0.008, *P* = 0.559), ASD^41^ (β = 7.20 × 10−5, *P* = 0.499), MDD^42^ (β = 0.051, *P* = 0.162), and insomnia^43^ (β = 0.041, *P* = 0.226; Table 2), indicating that our observed association between common genomic variations and gene expression differentiation are relatively specific to SCZ and bipolar disorder, possibly due to a high genetic overlap between the two disorders.^39^

We next focused on the set of 1,871 differentially expressed genes (DEGs) (adjusted *P* < 0.05, FDR corrected across 17,999 genes) identified by DESeq2. Out of the 1,871 DEGs, 416 genes overlapped with GWAS-derived SCZ risk genes (*P* = 6.15 ×10−5, hypergeometric testing) that were mapped from SCZ genomic loci by means of positional mapping, eQTL mapping, and 3D chromatin interaction mapping.^44^ For any of these three SNP-to-gene mapping approaches, the enrichment of DEGs in the GWAS-derived genes remained significant (Supplementary Table 2). Considering down-regulated (*n* = 1,268) and up-regulated DEGs (*n* = 603) separately revealed similar findings (Supplementary Table 3).

Blood-derived DEGs were also significantly enriched in DEGs revealed in previous transcriptome-wide association studies in SCZ^45^ (*P* = 0.022; Supplementary Table 4). Gene-set enrichment analysis through FUMA GENE2FUNC^44^ additionally showed significant enrichment of the DEGs in gene ontology (GO)^46^ cellular components of the whole membrane, neuron, endosome, et cetera (Supplementary Table 5), as well as in GWAS catalog reported gene-sets “Body mass index”, “Inflammatory bowel disease”, “Bipolar disorder”, “SC”, et cetera (Supplementary Table 6).

### Blood DEGs are associated with gene expression differentiation in the brain

We continued by examining how blood DEGs are associated with gene expressions in the brain. Using gene expression data from GTEx^47^ we found 1,838 out of the identified 1,871 DEG genes to show non-zero expression levels in all brain tissues,^16^ with a significant correlation observed between blood and brain cortical expression levels among these 1,838 genes (Spearman’s ρ = 0.430, *P* < 0.001; Supplementary Fig. 1), suggesting correlated gene expressions between blood and brain. Moreover, we examined to what extent DEGs observed in our blood samples also showed differentiated gene expression in brain samples of SCZ.^6^ Permutation analysis showed down-regulated DEGs (*n* = 1,268) to display significantly lower log2-FC (i.e. more negative) of expression levels in brain samples^6^ compared to null distributions of the log2-FC of randomly selected, same-sized gene sets (*P* = 0.016, 5,000 permutations). Up-regulated DEGs as observed in our blood samples (*n* = 603) were not found to show a significant effect compared to null distributions of log2-FC of random genes (*P* = 0.113, 5,000 permutations).

### Gene expression of DEGs covaries brain gray matter volume

Having established the linkage of DEGs with both genomic variations and transcriptions in the brain, we next examine the micro-macro association between inter-individual variations in gene expression of the 1,838 brain-expressed DEGs and brain gray matter volume (GMV). Using multivariate PLS analysis, we found a significant correlation between gene expression and GMV, indicated by the first PLS latent component (LC1) that accounted for 42.0% transcription-GMV covariance (*P* < 0.001, FDR corrected across the first 10 LCs accounting for 80% covariance; Fig. 2A). This LC described positive correlations between individual-level transcriptional composite scores and brain GMV composite scores within both SCZ (*r* = 0.741, *P* < 0.001) and HC (*r* = 0.435, *P* < 0.001) (Fig. 2A). Out of 1,838 DEGs, 1,183 DEGs (64.6%) significantly contributed to LC1 (*P*_fdr_ < 0.05), with 942 out of the 1,199 DEGs showing positive PLS loadings (which indicate positive contributions to the component) and 241 DEGs showing negative PLS loadings (which indicate negative contributions). Permutation testing using randomly selected DEGs indicated the significance of LC1 (*P* < 0.001, 5000 permutations), suggestive of the gene specificity of the observed association between GMV and SCZ DEGs.

**Figure 2.**
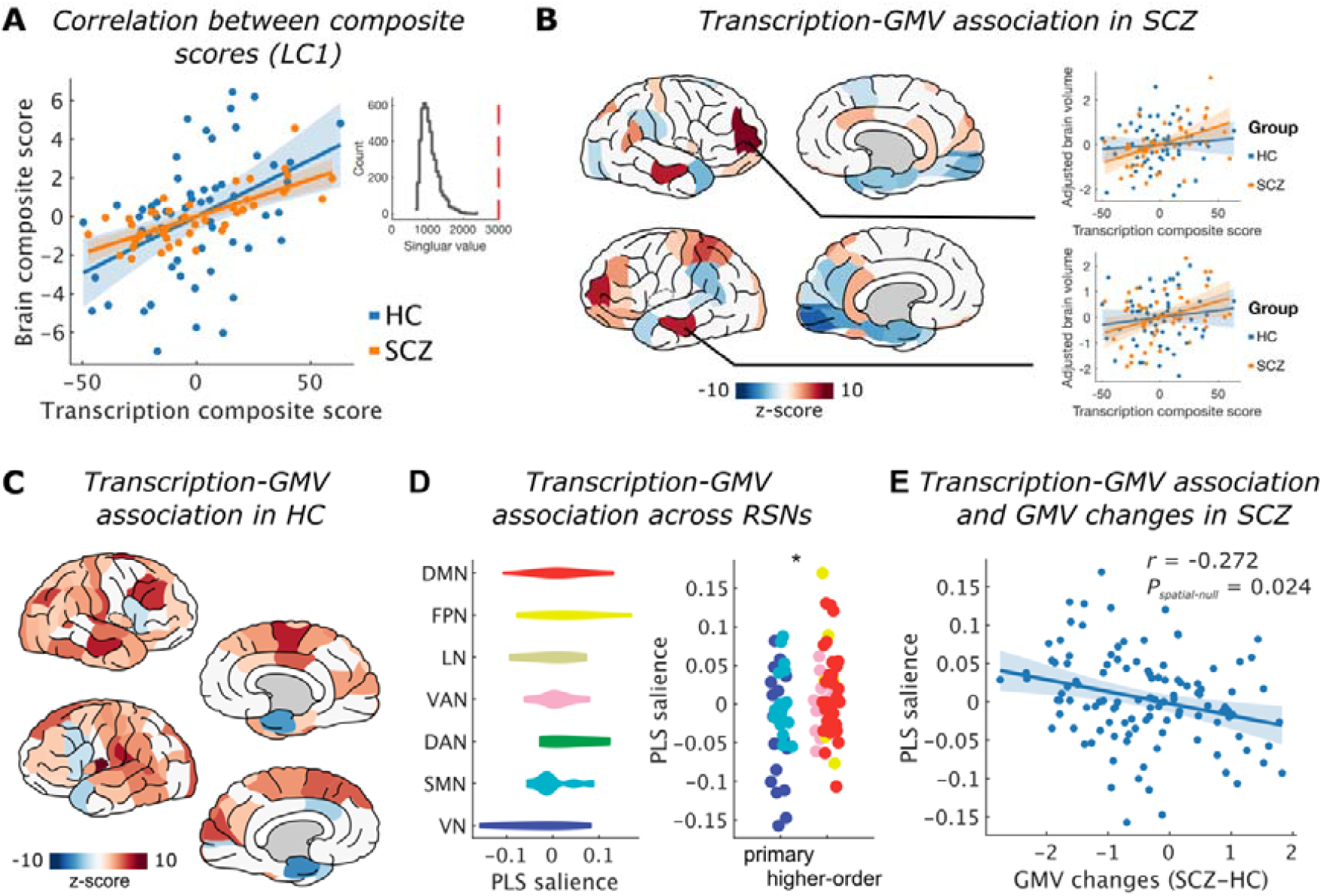
Association Between Gene Transcription and Brain Gray Matter Volume (GMV). **(A)** Gene transcription composite score significantly correlates to brain volume composite score within both SCZ (*r* = 0.749, *P* < 0.001) and HC (*r* = 0.435, *P* < 0.001) in the first latent component (LC1). **(B)** Regional contributions (*z*-scores) to the LC1 in SCZ. Red indicates positive associations between GMV and the transcription composite score; blue indicates negative associations. Two examples of the left middle temporal cortex and the right rostral middle frontal cortex are displayed. **(C)** Regional contributions (*z*-scores) to the LC1 in HC. **(D)** Within-SCZ transcription-GMV associations (described as PLS salience) across seven functional networks, including visual (VN), sensorimotor networks (SMN), dorsal attention network (DAN), ventral attention (VAN), limbic network (LN), frontoparietal (FPN), and default-mode networks (DMN). Higher-order cognitive networks (i.e., VAN, FPN, and DMN) show significantly larger PLS salience compared to primary VN and SMN (*t* = 2.688, *P* = 0.009). **(E)** The spatial correlation between transcription-GMV association (described as PLS salience) and GMV difference between SCZ and HC.

We further examined the contribution of each brain region to the multivariate transcription-GMV covariance as revealed in the PLS LC1. The transcriptional composite score was found to positively correlate to GMV across SCZ patients in bilateral rostral middle frontal, middle temporal, and superior parietal regions (adjusted *P* < 0.05, FDR corrected; Fig. 2B; Supplementary Table 5). In parallel, opposite correlations were found in bilateral pericalcarine, parahippocampal gyri, and entorhinal cortices (adjusted *P* < 0.05, FDR corrected). In HCs, the transcriptional composite score correlated to GMV of more distributed regions in bilateral temporal, inferior parietal, precentral/postcentral, and visual cortices across subjects (adjusted *P* < 0.05, FDR corrected; Fig. 2C).

Post-hoc analysis comparing the regional contributions to the transcription-GMV covariance in SCZ (i.e., PLS salience in LC1) in the context of the brain cognitive and primary networks^48^ revealed that regions of higher-order cognitive networks (i.e., DMN, FPN, and SN)^49^ tend to show significantly higher levels of contribution compared to regions of primary visual and sensorimotor networks (*t* = 2.688, *P* = 0.009; Fig. 2C). No effect was observed in HC (*t* = −1.179, *P* = 0.241). Furthermore, the spatial pattern of regional contributions to the transcription-GMV covariance in LC1 significantly correlated to the spatial pattern of GMV differences between SCZ and HC (*r* = −0.272, *P*_spatial-null_ = 0.012; permutation testing using the spatial null model that conserves spatial auto-correlation)^50^ (Fig. 2D), suggesting that the transcription-GMV covariance in SCZ also paralleled with the extent of brain volume alterations in SCZ. No effect was observed in HC, either (*r* = 0.080, *P*_spatial-null_ = 0.562).

### DEG-GMV covariance shows similar transcriptional profiles with SCZ genes

The spatial pattern of the transcription-GMV covariance in above LC1 was further correlated to whole-brain gene expression profiles of 15,071 genes from the Allen Human Brain Atlas (see details in Supplementary Methods). The top 500 genes with the highest correlations involved 118 genes that have been revealed in the previous GWAS on SCZ,^2^ which exceeded what could be expected by chance (hypergeometric testing: *P* = 0.016). Using surrogate brain maps conserving spatial auto-correlation^16,51^ (16,51) further confirmed this enrichment (*P* = 0.026, 1,000 permutations). Similar results were also observed for the top 200 (*P* = 0.033) and 1000 genes (*P* = 0.019). No significant enrichment could be observed in the set of 1,838 DEGs (*P* = .363).

### Gene expression of DEGs covaries brain connectivity of the sensorimotor network

Considering the associated spatial patterns of transcriptional profiles SCZ genes and brain disconnectivity,^14^ we further examined whether individual variations in gene expression also paralleled variations in brain connectivity. PLS analysis revealed a significant correlation (LC1: *P* < 0.001, 5,000 permutations; Fig. 3), where positive correlations were observed between transcriptional composite scores and the regional SC strength of the bilateral precentral and postcentral gyrus, the left superior parietal gyrus, posterior cingulate gyrus, et cetera, in the SCZ group (adjusted *P* < 0.05, FDR corrected; Fig. 3). Post-hoc examinations on the spatial distribution of the regional contributions (i.e., PLS salience) across resting-state networks showed the highest contributions in regions of the sensorimotor network (SMN) (Fig. 3) (SMN vs. the rest: *t* = 4.347, adjusted *P* < 0.001, FDR corrected). Significant correlations of the SMN were also observed in the PLS analyses for FC and were reported in Supplementary Results and Supplementary Fig. 2.

**Figure 3.**
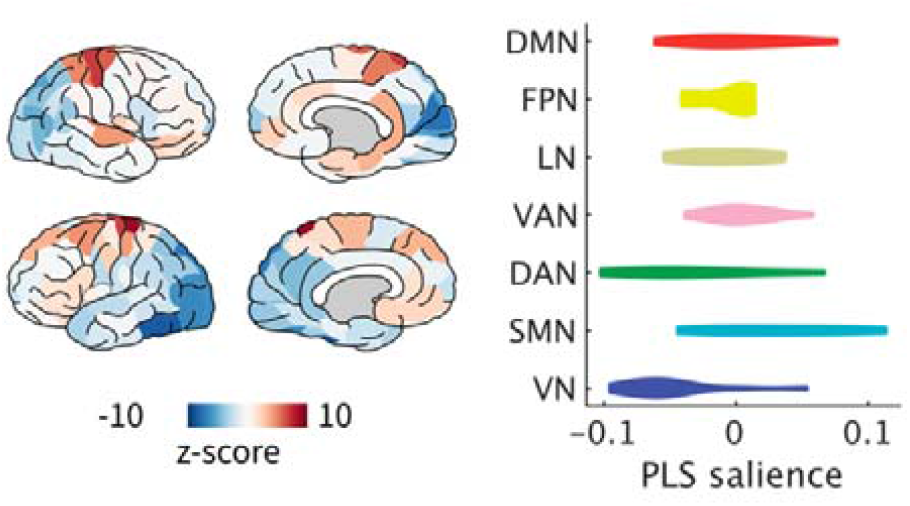
Association among Transcription, GMV, and Brain Connectivity. Left: Correlations of structural connectivity (SC) strength with the transcriptional composite score revealed in the PLS analysis within SCZ. Red indicates positive correlations; blue indicates negative correlations. Right: within-SCZ transcription-SC correlations across seven functional networks.

**Figure 4.**
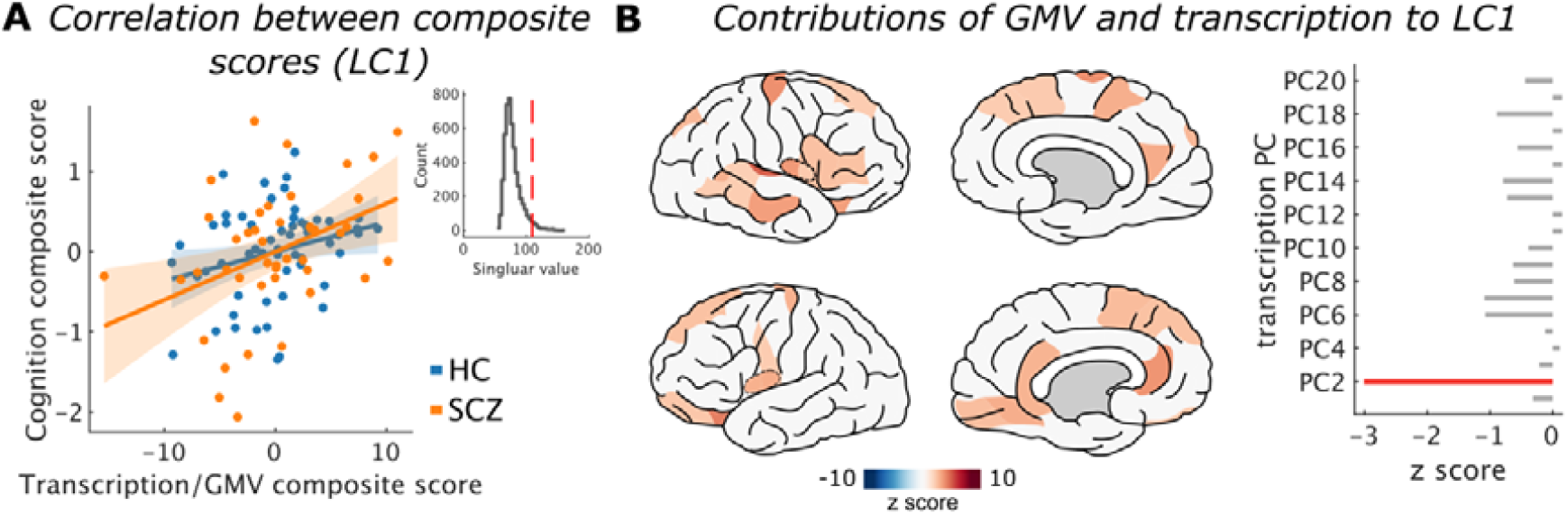
Association among Transcription, GMV, and Performance in the WAIS test. **(A)** GMV and transcription composite score significantly correlates to cognition composite score within both SCZ (*r* = 0.396, *P* = 0.008) and HC (*r* = 0.267, *P* = 0.044) in the first latent component (LC1). **(B)** Regional GMV (left) and transcription PCs’ (right) contributions (*z*-scores) to the LC1. Positive *z* score indicates a positive association with the performance in the WAIS test.

### Associations among transcription, brain structure, and clinical characteristics

To examine how cross-scale biological features together influence clinical characteristics in SCZ, we performed PLS analysis to examine associations among transcription, gray matter volume, and clinical symptoms and cognitive functions in SCZ patients. Here, twenty principal components (PCs) of the transcriptional data (explained variance: 95.7%) were included in the PLS analysis. A significant LC was observed (*P* = 0.031, 5,000 permutations), showing significantly positive correlations between individuals’ GMV and cognitive performance in the WAIS test (*z* > 2.595, adjusted *P* < 0.05; Fig. 3A). This correlation was driven by GMV in bilateral superior frontal regions, insula, precuneus, left inferior frontal regions, right orbital frontal regions, et cetera (*q* < 0.05, FDR corrected; Fig. 3B), as well as the second transcriptional PC (*z* = −2.952, adjusted *P* = 0.019). No significant effect was observed for the PANSS score (*P* > .109).

### Split-half validation shows robust correlation patterns

To examine the robustness of the observed association between gene expression and GMV, we randomly divided data samples into two halves and repeated the PLS correlation analyses in both sub-samples. Both halves showed significant LC1 (*P*s < 0.001, 5,000 permutations, FDR corrected), confirming associated DEGs and GMV. In SCZ, 43 regions were found to significantly contribute to LC1 in the first half and 37 regions were found in the second half (Supplementary Results and Supplementary Fig. 3). The spatial pattern of regional contributions in the two halves significantly correlated to each other (Pearson’s *r* = 0.42, *P* < 0.001), suggesting the robustness of the association between gene expression and GMV.

## Discussion

The integration between MRI and transcriptomics has established the transcriptomic correlates of cerebral alterations in relation to SCZ.^52,53^ Here we present a multi-omic study in SCZ, examining blood transcriptomic data, neuroimaging data, and data on clinical symptoms and cognitive impairments. Individual variations of macroscale brain volume in SCZ are associated with transcriptional differentiations, which correlate to genomic variations of SCZ, accounting for transcriptomic contributions to brain structure and function in SCZ. Furthermore, brain volume and transcription show a correlation to the general cognitive abilities. Our study suggests that blood-sample gene expressions might provide an informative, *in vivo* way to study participant-level transcriptomic-neuroimaging associations in SCZ.

Cross-scale associations between gene expression and brain gray matter show that lower expression of DEGs in SCZ are associated with reduction in gray matter volume of frontotemporal regions. The involvement of frontal and temporal brain regions in SCZ has been broadly reported in previous studies, with decreased gray matter, surface area, and cortical thickness observed in SCZ patients compared to HC.^54^ On the other side, transcriptome studies examining post-mortem brain materials have noted downregulation of immune-associated genes in patients.^55^ Our reported transcriptome-neuroimaging associations at the participant level further corroborate on recent findings linking the transcriptome and the brain on the basis of brain spatial variations,^12-14,56^ for instance, genes related to the central nervous system development overlap with those related to brain volume changes in SCZ.^12^ Associations between transcription and brain volume could be attributed to the crucial regulatory role of SCZ risk genes in human brain development.^57^ In line with this, neuron- and astrocyte/oligodendrocyte progenitors-linked genes are known to play an important role in transcriptomic and polygenic manifestations of cortical thickness heterogeneity in SCZ.^58^ The observed participant-level relationships between gene transcription and brain volume in SCZ shows great potential for integrating multi-omics features in future works on predictive models in SCZ.

The structural and functional connectivity of the somatomotor network tend to be associated with gene transcription and gray matter volume. The somatomotor network has been frequently reported in previous neuroimaging studies, showing differentiated functional connectivity in SCZ compared with HCs.^59,60^ Our reported associations are also in line with previous studies showing structural connectivity to be related to a tight coupling of transcriptional profiles.^61^ SCZ risk genes specifically showed a brain transcriptional profile that overlapped with the spatial pattern of disruptions in white matter connectivity.^14^ Such an association between SCZ gene expression and brain dysconnectivity might reflect the genetic origins of widespread connectivity disruptions observed in SCZ.^10,25,62,63^

The cognitive impairments of patients were found to be associated with individual variations in transcriptome and neuroimaging measures, predominantly involving areas of the default mode network (precuneus, middle temporal gyrus, and prefrontal lobe). This result extended the relationship between brain structures in cognitive networks and cognitive domains in SCZ,^64^ showing an involvement of genetic component in this relationship. Such associations across transcription, brain, and cognition are also compatible with the shared genetic loci between intelligence and the disorder, with poorer cognitive performance reported to be linked to most SCZ risk alleles^65^ and a higher SCZ polygenic risk score.^10,66^ Further, cognitive dysfunction in SCZ patients has been related to altered expression of immunological genes as measured in blood samples.^67^ We add to this by providing an empirical evidence for the association across cognitive abilities, GMV, and gene expression. Integrating genetics with neuroimaging is key to identify novel genetic markers of SCZ and improve the predictability of the disorder.^68^

Our reported relationship between DEGs in blood samples of SCZ and common variants and brain expression differentiation in SCZ implies blood transcriptomic data to be an informative source to study transcriptome-neuroimaging associations in SCZ. Genetic association between SCZ and cerebral morphometry are well demonstrated (Psychiatric Genomics Consortium, CLOZUK, and UK Biobank),^69^ but one of the obstacles in identifying transcriptome-neuroimaging associations of the disorder is that subsequent analysis of transcriptome and MRI data are commonly performed on data coming from different sources. Our study now attempts to address this issue by analyzing cross-scale data from one cohort of SCZ. Our observed correspondence between blood DEGs and brain DEGs is in line with the correlation of genetic effects between human blood and brain.^17,18^ Our results of associations between blood DEGs and SCZ GWAS further suggest blood DEGs to be significantly regulated by common genomic variants in SCZ, regardless of the examined ethnics. There were 11 overlapping genes out of the 61 genes reported in a SCZ transcriptome-wide association study,^45^ uncovering a specific set of genes that may represent an authentic susceptibility gene for the disease.^65^ Current knowledge suggests that the cause of SCZ is highly heterogeneous.^70^ Therefore, examinations of the participant-level associations between blood gene expression and imaging-derived brain phenotypes in SCZ might provide evidence of disease-involved biological pathways, advancing our understanding of the disease mechanisms.

A first limitation of the current study is the gap between transcriptomes derived from intravenous blood and brain tissue. Although we showed the relationship between blood DEG and brain DEG, they are not equivalent. Obtaining gene expression levels of brain tissue in vivo is, however, almost impossible for SCZ patients. Second, we were unable to evaluate the effects of cumulative exposure to antipsychotics. Third, in our study brain phenotypes were only acquired on the basis of structural MRI sequences. Combining structural MRI with positron emission tomography revealed a link between hippocampal morphological alterations and microglial activation in first-episode psychosis patients,^71^ showing the potential of combining multiple modalities of brain imaging in SCZ studies. Large collaborative longitudinal multimodal imaging studies of SCZ will be required to clarify the neurodevelopmental trajectory and specific medication influences.

In conclusion, we show that individual variations in gene expression are related to variations in brain structure in SCZ, providing an informed neurogenetic profile of the disorder. Establishing transcriptomic correlates of brain structure holds promise for building biomarkers that could improve the diagnosis and assessment of SCZ.^68^ Understanding the underlying multi-scale mechanisms might also build up a plausible path toward developing novel treatments.^72^ Reflecting the complex and multifactorial nature, micro- and macro-scale neurobiology implicated in SCZ provide leads for clinical translation.

## Data Availability

All data produced in the present study are available upon reasonable request to the authors.

## Supplementary material

Supplementary material is available at Brain online.

