## Supplementary material for "Multi-omic transcriptional, brain, and clinical variations in schizophrenia": Tables

**Table 1 Demographical and Clinical Data of Participants**

| **Characteristic** | **Patients (n = 43)** | **HCs (n = 60)** | ***P* value** |
| --- | --- | --- | --- |
| **Sociodemographic** |  |  |  |
| Age, y | 26.5 ± 9.4 | 26.0 ± 5.5 | .783^a^ |
| Gender, male/female | 23/20 | 36/24 | .510^b^ |
| Ethnicity, Han/others | 43/0 | 48/0 |  |
| Education level, y | 12.7 ± 2.9 | 14.0 ± 2.9 | .078^c^ |
| **Clinical assessment^d^** | | |  |
| PANSS total score | 61.7 ± 17.4 | NA |  |
| PANSS positive score | 13.6 ± 6.5 | NA |  |
| PANSS negative score | 14.8 ± 8.3 | NA |  |
| PANSS general psychopathology score | 33.3 ± 8.3 | NA |  |
| **Intelligence^c,e^** |  |  |  |
| Information | 9.5 ± 3.1 | 12.7 ± 3.2 | 2.381 × 10^-6^ |
| Digit span | 11.5 ± 2.6 | 14.1 ± 3.2 | 8.905 × 10^-6^ |
| Digit symbol coding | 11.0 ± 2.7 | 15.6 ± 3.0 | 7.090 × 10^-11^ |
| Vocabulary | 10.8 ± 2.6 | 13.6 ± 3.0 | 1.088 × 10^-6^ |
| Picture completion | 8.3 ± 2.8 | 11.1 ± 3.0 | 2.146 × 10^-6^ |
| Block design | 10.6 ± 3.3 | 13.9 ± 4.0 | 1.254 × 10^-6^ |
| IQ | 97.6 ± 14.6 | 114.7 ± 17.5 | 7.312 × 10^-7^ |

Note: HCs, healthy controls; NA, not applicable; PANSS, Positive and Negative Syndrome Scale; IQ, Intelligence quotient.

^a^Two sample t-test.

^b^χ^2^ test.

^c^Wilcoxon Rank Sum test.

^d^Seven patients had missing PANSS information.

^e^One healthy control did not take the intelligence test. Information and vocabulary were unavailable for one healthy control.

**Table 2 Associations Between Gene Expression Differentiation and Genomic Variations**

| **GWAS** | **Reference** | ***β*** | **SE** | ***P* value** |
| --- | --- | --- | --- | --- |
| SCZ (PGC wav3) | Trubetskoy et al.(2) | 0.196 | 0.066 | 0.002* |
| SCZ (East Asian) | Lam et al.(36) | 0.135 | 0.056 | 0.008* |
| BD | Mullins et al.(38) | 0.165 | 0.021 | 0.003* |
| ADHD | Demontis et al.(40) | -0.008 | 0.055 | 0.56 |
| ASD | Grove et al.(41) | 7.20×10^-5^ | 0.053 | 0.499 |
| MDD | Wray et al.(42) | 0.051 | 0.051 | 0.612 |
| Insomnia | Jansen et al.(43) | 0.041 | 0.055 | 0.226 |

Note: ADHD, attention-deficit/hyperactivity disorder; ASD, autism spectrum disorder; BD, bipolar disorder; MDD, major depressive disorder; SCZ, schizophrenia.

*Significant (adjusted P < 0.05, FDR corrected).
