## Supplementary material for "Multi-omic transcriptional, brain, and clinical variations in schizophrenia": SI

**Content**

### Supplemental Methods

#### Inclusion and exclusion criteria

From April 2020 to September 2020, 43 patients with schizophrenia were recruited from the Department of Psychiatry at Xijing Hospital. There were also 60 matched healthy controls, who were enrolled through advertising. The participants were diagnosed on the basis of the Diagnostic and Statistical Manual of Mental Disorders, Fifth Edition (DSM-5), and consensus diagnoses were made by two experienced clinical psychiatrists using all the available information. They were all right-handed, and their biological parents were of the Han Chinese ethnic group. The exclusion criteria for patients were as follows: (1) the presence of another psychiatric disorder; (2) a history of repetitive transcranial magnetic stimulation, transcranial current stimulation, or behavioral treatment; (3) a history of clinically significant neurological, neurosurgical, or medical illnesses; (4) substance abuse within the prior 30 days or substance dependence within the prior 6 months; (5) pregnancy or any other MRI contraindications, e.g., cardiac pacemakers and other metallic implants; (6) unwillingness to undertake the scanning. Exclusion criteria for healthy controls were as follows: (1) the presence of any psychotic syndrome; (2) a history of receiving antipsychotics, repetitive transcranial magnetic stimulation, transcranial current stimulation, or behavioral treatment; the remaining (3), (4), (5), and (6) were the same as the exclusion criteria for patients.

#### RNA-seq data collection and analysis

Total RNA was extracted using Trizol reagent kit (Invitrogen, Carlsbad, CA, USA). RNA quality was assessed on an Agilent 2100 Bioanalyzer (Agilent Technologies, Palo Alto, CA, USA) and checked using RNase free agarose gel electrophoresis. After total RNA was extracted, eukaryotic mRNA was enriched by Oligo(dT) beads, while prokaryotic mRNA was enriched by removing rRNA by Ribo-ZeroTM Magnetic Kit (Epicentre, Madison, WI, USA). Then the enriched mRNA was fragmented into short fragments using fragmentation buffer and reverse transcripted into cDNA with random primers. Second-strand cDNA were synthesized by DNA polymerase I, RNase H, dNTP and buffer. Then the cDNA fragments were purified with QiaQuick PCR extraction kit (Qiagen, Venlo, The Netherlands), end repaired, poly(A) added, and ligated to Illumina sequencing adapters. The ligation products were size selected by agarose gel electrophoresis, PCR amplified, and sequenced using Illumina HiSeq2500 by Gene Denovo Biotechnology Co. (Guangzhou, China).

Reads resulted from the sequencing machines were raw reads containing adapters or bases with low quality. Thus, to get high-quality clean reads, reads were further filtered by fastp^1^ (version 0.18.0). The parameters include: 1) removing reads containing adapters; 2) removing reads containing more than 10% of unknown nucleotides (N); 3) removing low quality reads containing more than 50% of low quality (Q-value ≤ 20) bases. An index of the reference genome was built, and paired-end clean reads were mapped to the reference genome using HISAT2. 2.4^2^ with “-rna-strandness RF” and other parameters set as default.

#### Image acquisition

MRI data were acquired using a General Electric (GE) Discovery MR750 3.0 T scanner at the Department of Radiology at Xijing Hospital with a standard 8-channel head coil. Participants were instructed to relax with their eyes closed but keep from falling asleep during their MRI scan. Scanning parameters are tabulated in Supplemental Table S1.

#### DWI data preprocessing

DWI data preprocessing included: 1) realignment of the 64 diffusion-weighted volumes (b = 1000 s/mm^2^) and the b = 0 volume and corrections for small head movements and gradient-induced distortions;^3^ 2) reconstruction of voxel-wise diffusion peaks within the brain mask using Constrained Spherical Deconvolution (CSD);^4^ 3) reconstruction of fiber tracts through deterministic streamline tractography using Fiber Assignment by Continuous Tracking (FACT) adjusted for the use of multiple diffusion peaks.^5^ Eight streamline seeds were started for each brain voxel and tracking was stopped if a streamline reached a voxel of low preferred diffusion direction (FA < 0.1), exceeded the brain mask, and/or made a sharp turn (> 45°).

#### Functional MRI data preprocessing

Resting-state functional MRI (fMRI) data of each subject were realigned and co-registered with the T1-weighted image to overlap with the resultant cortical parcellation maps. Next, the blood oxygenation level-dependent (BOLD) time series were corrected for linear trends, as well as global nuisance covariance, including 6 head motion parameters and mean signals of white matter and cerebrospinal fluid. Third, band-pass filtering (0.01-0.1 Hz) was performed together with motion scrubbing.^6^ Each slice with framewise displacement (FD) exceeding 0.25 (defined as the sum of the absolute derivatives of the six realignment parameters) and dynamic variability (DVARS) exceeding 1.5, as well as 1 back neighbor, were removed.^6^

#### Permutation testing on the correspondence of blood and brain DEGs

Permutation testing was performed to examine whether the observed DEGs in blood samples also showed differentiated expression in brain samples in SCZ. Differential gene expressions (log2-FC) through microarray meta-analysis based on brain samples of SCZ were obtained from^7^. The mean log2-FC was computed for the set of DEGs identified in blood samples and was compared to a null distribution of mean log2-FCs obtained for the same-sized random gene sets. A two-sided *P* value was assigned according to the proportion of the null distribution that exceeds the mean log2-FC of the blood DEGs.

#### PLS analysis

Permutation testing was used to statistically evaluate the observed correlation, with an LC identified as significant when the singular value exceeded a computed null distribution of singular values obtained from PLS analyses on random permutations of subjects (5,000 permutations). For any LC, the individual-level composite scores^8^ for gene expressions and imaging-derived phenotypes were computed, separately, to assess the level of covariance captured by the LC.

Bootstrapping (with 5,000 samplings) was further used to estimate whether the resulting PLS saliences of the significant LCs were robust.^9^ Bootstrap ratio (BSR) was calculated by dividing the PLS salience by the standard error estimated in the bootstrapping. BSR values were equivalent to Z-scores as the bootstrap distribution followed a normal distribution.^10^ *P* values were obtained according to the Z-score, and significant genes and brain metrics were identified subsequently (corrected for multiple comparisons using the Bonferroni correction, *α* < 0.05). PLS analysis was conducted using myPLS (<https://github.com/danizoeller/myPLS>).^9^ For validation purposes, the dataset was randomly split into two halves, with PLS analysis separately conducted in each half (Supplementary Results).

### Supplemental Results

#### Gene expression, GMV, and FC

PLS analysis examining correlations across gene expressions, gray matter volume, and FC strength revealed a significant LC1 (*P* < .001, 5,000 permutations; **Supplemental** **Figure S2**), showing robust positive associations in the bilateral orbital and inferior frontal cortex, left fusiform, anterior cingulate gyrus, et cetera, in the SCZ group (*adjusted P < 0.05, FDR corrected;* **Supplemental Figure S2B**). Across functional networks, regions of the default-mode network showed trend-level, non-significant higher contributions compared to the rest of the brain (*t* = 2.213, uncorrected *P* = .029; *P*_fdr_ = .101), while regions of the SMN showed significantly lower contributions (*t* = −6.487, *P*_fdr_ < .001).

#### Validation using the DK-68 atlas

To investigate whether the selection of the brain atlas affects our main results, we redid PLS analyses based on the Desikan-Killiany Atlas^12^ that divides the brain into 68 brain regions. This analysis again revealed a significant latent component that accounted for 41.9% transcription-volume covariance (LC1, *P* < .001, FDR corrected across the first 10 components; **Supplemental Figure S4A**), showing a relationship between individual-level transcriptional composite scores and brain gray matter volume composite scores within both SCZ (*r* = 0.668, *P* < .001) and HC (*r* = 0.412, *P* < .001). Out of 1,838 DEGs, 1,133 DEGs (63.4%) significantly contributed to LC1 (*P* < .05, FDR corrected), with 899 out of the 1,133 DEGs showing positive PLS loadings and 234 DEGs showing negative PLS loadings. Considering contributions of each brain region to the transcription-GMV covariance captured in LC1, we noted a similar pattern as the main results: transcriptional composite score positively correlated to brain volume in bilateral rostral middle frontal, middle temporal, and superior parietal regions across SCZ patients (adjusted *P* < .05, FDR corrected; **Supplemental Figure S4B**). These results thus suggested that the choice of the Desikan-Killiany atlas results in similar findings as our main results.

#### Split-half validations

We examined the robustness of the association between gene expressions and brain volume by randomly splitting data samples into two halves and repeating the PLS correlation analyses in both sub-samples. The two halves respectively included 52 subjects (26 controls and 26 patients; referred to as H1) and 51 subjects (32 controls and 19 patients; referred to as H2). The first LC appeared to be significant in both H1 (*P* < .001, 5,000 permutations, FDR corrected) and H2 (*P* < .001, 5,000 permutations, FDR corrected), suggestive of associated gene expressions and gray matter volume. The LC1 was significantly contributed by 1,290 genes in H1 and 991 genes in H2, in which 1,077 (83.5%) and 782 (78.9%) genes showing a high overlap with the genes reported in the main results. In SCZ, 43 regions were found to significantly contribute to LC1 in H1 (30 out of 43 reported in the main results; **Supplemental Figure S3A**) and 37 regions were found in H2 (25 out of 43 reported in the main results; **Supplemental Figure S3B**). The spatial pattern of regional contributions (i.e. PLS salience) in H1 and H2 significantly correlated to each other (Pearson’s *r* = 0.42, *P* < .001; **Supplemental Figure S3C**), suggesting that the association between gene expression and gray matter volume tends to be robust in the split-half analyses.

### Supplemental Figures

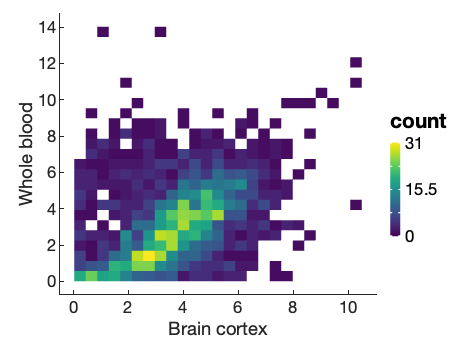

**Figure S1***.* Gene expressions in the brain cortex and the whole blood in the GTEx dataset.

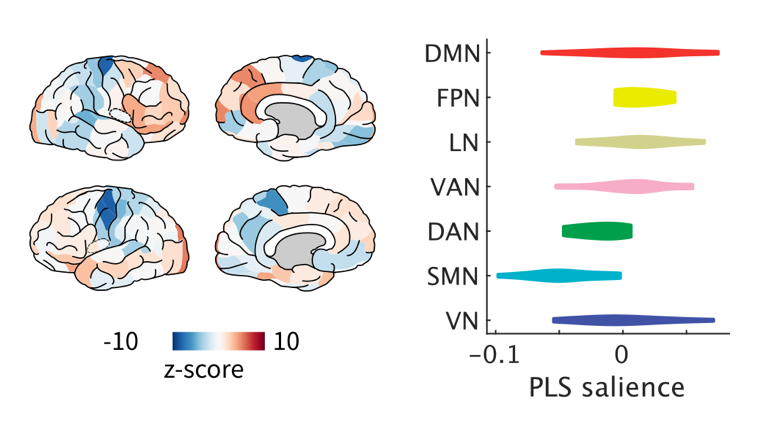

**Figure S2*.*** *Association Among Gene Transcription, Gray Matter Volume, and Functional Connectivity (FC)****.*** Left: Correlations of FC strength with the transcriptional composite score within SCZ. Right: within-SCZ transcription-FC correlations across seven functional networks.

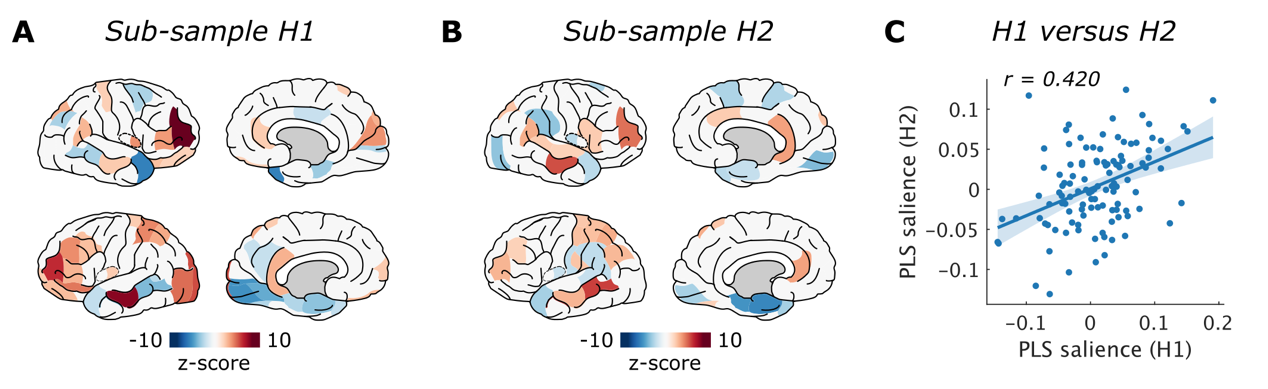

**Figure S3***. Split-half validation analysis.* (**A**) Brain plots of regions showing significant contributions to the transcription-GMV association captured in the LC1 in SCZ, based on the sub-sample of the first half (H1). (**B**) Brain plots of regions showing significant contributions to the transcription-GMV association captured in the LC1 in SCZ, based on the sub-sample of the second half (H2). (**C**) Regional contributions to the LC1 (i.e. PLS salience) in H1 are correlated to those in H2 (*r* = 0.420, *P* < .001).

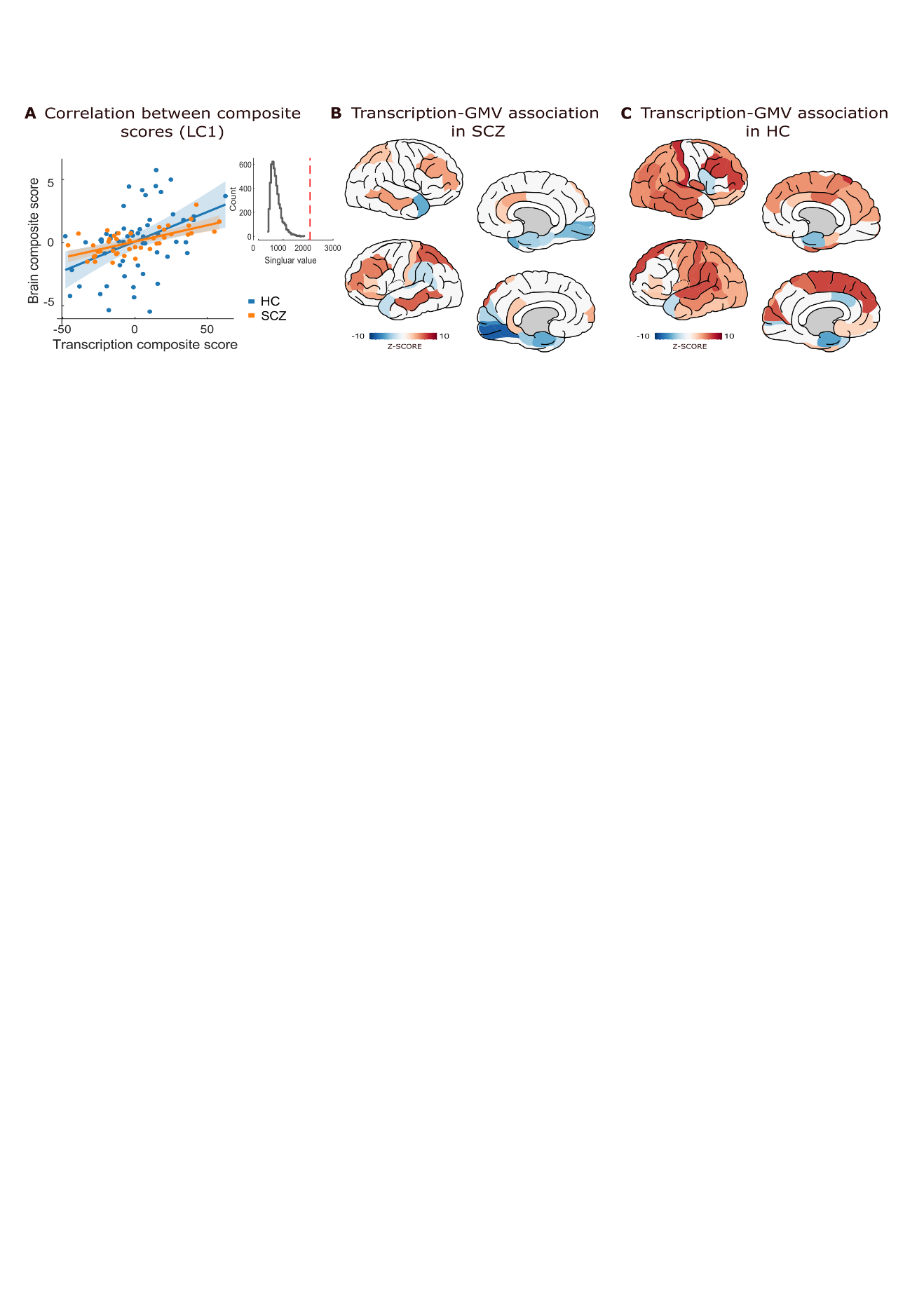

**Figure S4***. Transcription-GMV association based on the Desikan-Killiany atlas.* (**A**) Gene transcription composite score significantly correlates to brain volume composite score based on Desikan-Killiany atlas within both SCZ (*r* = 0.668, *P* < .001) and HC (*r* = 0.412, *P* < .001) in the first latent component (LC1). (**B**) Regional contributions (z-scores) to the LC1 in SCZ (adjusted *P* < .05, FDR corrected). (**C**) Regional contributions (z-scores) to the LC1 in HC (adjusted P < .05, FDR corrected). Red indicates positive associations between GMV and the transcription composite score; blue indicates negative associations.

### Supplemental Tables

**Table S1. Scanning parameters**

|  | T1-weighted imaging | Diffusion weighted imaging | Functional imaging |
| --- | --- | --- | --- |
| TR (ms) | 8.2 | 10000 | 2000 |
| TE (ms) | 3.2 | 82.4 | 30 |
| Flip angle (◦) | 12 | NA | 90 |
| Field of view (mm^2^) | 256 × 256 | 240 × 240 | 240 × 240 |
| Matrix | 256 × 256 | 128 × 128 | 64 × 64 |
| Slice thickness (mm) | 1 | 2 | 3.5 |
| Slice gap (mm) | 0 | 0 | 0 |
| Number of slices | 196 | 70 | 45 |

NA, not applicable; TE, echo time; TR, repetion time.

**Table S2. Overlaps between DEGs and risk genes derived from GWAS using different snp-to-gene mapping approaches**

|  | N | n | P value | Ajusted P value |
| --- | --- | --- | --- | --- |
| scz_all | 3379 | 416 | 3.073E-05 | 6.147E-05 |
| scz_pos | 769 | 103 | 2.941E-03 | 2.941E-03 |
| scz_eqtl | 1803 | 243 | 5.127E-06 | 2.051E-05 |
| scz_ci | 2783 | 344 | 1.279E-04 | 1.705E-04 |

N: the number of genes mapped from snps. n: the number of overlapping genes.

**Table S3. Overlaps between up-regulated/down-regulated DEGs and risk genes derived from GWAS**

|  | N | n | P value | Ajusted P value |
| --- | --- | --- | --- | --- |
| down-regulated | 3379 | 279 | 1.185E-03 | 2.369E-03 |
| up-regulated | 3379 | 137 | 5.787E-03 | 5.787E-03 |

**Table S4. Overlaps between DEGs and risk genes reported in TWAS on SCZ**

| Ensemble ID | Symbol | log-FC | Ajusted P value |
| --- | --- | --- | --- |
| ENSG00000079313 | REXO1 | -0.500 | 0.002 |
| ENSG00000101445 | PPP1R16B | -0.429 | 0.002 |
| ENSG00000137500 | CCDC90B | 0.222 | 0.005 |
| ENSG00000139970 | RTN1 | -0.305 | 0.026 |
| ENSG00000149930 | TAOK2 | -0.433 | 0.000 |
| ENSG00000154822 | PLCL2 | 0.245 | 0.009 |
| ENSG00000158792 | SPATA2L | -0.382 | 0.014 |
| ENSG00000170571 | EMB | 0.158 | 0.041 |
| ENSG00000175213 | ZNF408 | -0.303 | 0.013 |
| ENSG00000262246 | CORO7 | -0.426 | 0.002 |
| ENSG00000266472 | MRPS21 | 0.374 | 0.031 |

**Table S5. Gene-set analysis for gene sets involved in GO cellular components**

| GeneSet | N | n | adjusted P |
| --- | --- | --- | --- |
| GO_NUCLEOPLASM_PART | 1113 | 188 | 4.300E-44 |
| GO_NUCLEAR_BODY | 769 | 135 | 5.500E-33 |
| GO_WHOLE_MEMBRANE | 1647 | 207 | 1.010E-29 |
| GO_CYTOPLASMIC_VESICLE_PART | 1483 | 188 | 2.360E-27 |
| GO_CYTOSKELETAL_PART | 1639 | 198 | 2.800E-26 |
| GO_CATALYTIC_COMPLEX | 1351 | 169 | 6.770E-24 |
| GO_MICROTUBULE_CYTOSKELETON | 1169 | 153 | 1.430E-23 |
| GO_GOLGI_APPARATUS | 1547 | 183 | 2.970E-23 |
| GO_CHROMOSOME | 1059 | 140 | 5.660E-22 |
| GO_CHROMATIN | 557 | 94 | 7.630E-22 |
| GO_NUCLEAR_SPECK | 389 | 74 | 3.260E-20 |
| GO_VACUOLE | 760 | 109 | 9.530E-20 |
| GO_ENDOSOME | 885 | 120 | 9.530E-20 |
| GO_ENDOPLASMIC_RETICULUM | 1884 | 197 | 5.400E-19 |
| GO_TRANSFERASE_COMPLEX | 772 | 108 | 8.730E-19 |
| GO_ACTIN_CYTOSKELETON | 491 | 81 | 2.710E-18 |
| GO_NUCLEAR_OUTER_MEMBRANE_ENDOPLASMIC_RETICULUM_MEMBRANE_NETWORK | 1080 | 131 | 1.710E-17 |
| GO_CELL_JUNCTION | 1275 | 145 | 6.390E-17 |
| GO_GOLGI_MEMBRANE | 747 | 101 | 1.420E-16 |
| GO_ENDOPLASMIC_RETICULUM_PART | 1344 | 147 | 1.040E-15 |
| GO_NUCLEAR_CHROMATIN | 369 | 64 | 1.410E-15 |
| GO_MICROTUBULE_ORGANIZING_CENTER | 722 | 96 | 2.600E-15 |
| GO_GOLGI_APPARATUS_PART | 964 | 115 | 7.190E-15 |
| GO_ANCHORING_JUNCTION | 552 | 80 | 7.560E-15 |
| GO_NUCLEAR_CHROMOSOME | 599 | 83 | 2.820E-14 |
| GO_VESICLE_MEMBRANE | 780 | 97 | 1.170E-13 |
| GO_SECRETORY_GRANULE | 831 | 100 | 3.500E-13 |
| GO_NEURON_PART | 1709 | 165 | 7.150E-13 |
| GO_CENTROSOME | 500 | 71 | 8.920E-13 |
| GO_MITOCHONDRION | 1552 | 153 | 1.180E-12 |
| GO_SECRETORY_VESICLE | 983 | 110 | 1.930E-12 |
| GO_CELL_PROJECTION_PART | 1438 | 144 | 1.950E-12 |
| GO_VACUOLAR_PART | 553 | 74 | 4.860E-12 |
| GO_PERINUCLEAR_REGION_OF_CYTOPLASM | 697 | 85 | 1.410E-11 |
| GO_CELL_SUBSTRATE_JUNCTION | 409 | 60 | 1.810E-11 |
| GO_POLYMERIC_CYTOSKELETAL_FIBER | 706 | 83 | 1.680E-10 |
| GO_CELL_LEADING_EDGE | 397 | 56 | 4.500E-10 |
| GO_ENDOSOMAL_PART | 514 | 66 | 5.090E-10 |
| GO_VACUOLAR_MEMBRANE | 399 | 56 | 5.240E-10 |
| GO_NEURON_PROJECTION | 1301 | 126 | 5.480E-10 |
| GO_ORGANELLE_SUBCOMPARTMENT | 375 | 52 | 4.010E-09 |
| GO_MICROTUBULE | 410 | 55 | 4.190E-09 |
| GO_CELL_CELL_JUNCTION | 447 | 58 | 4.820E-09 |
| GO_ENVELOPE | 1166 | 113 | 5.250E-09 |
| GO_MITOCHONDRIAL_PART | 1019 | 102 | 6.520E-09 |
| GO_PLASMA_MEMBRANE_REGION | 1185 | 113 | 1.250E-08 |
| GO_SUPRAMOLECULAR_COMPLEX | 936 | 94 | 2.510E-08 |
| GO_SOMATODENDRITIC_COMPARTMENT | 818 | 85 | 3.070E-08 |
| GO_SYNAPSE | 1169 | 109 | 7.510E-08 |
| GO_MEMBRANE_PROTEIN_COMPLEX | 1153 | 106 | 2.350E-07 |
| GO_CYTOPLASMIC_REGION | 488 | 57 | 2.350E-07 |
| GO_MITOCHONDRIAL_ENVELOPE | 725 | 73 | 1.190E-06 |
| GO_DENDRITIC_TREE | 588 | 60 | 9.430E-06 |
| GO_SYNAPSE_PART | 932 | 83 | 2.210E-05 |
| GO_CELL_BODY | 557 | 56 | 3.050E-05 |
| GO_POSTSYNAPSE | 610 | 57 | 1.670E-04 |
| GO_AXON | 600 | 56 | 2.000E-04 |
| GO_NUCLEOLUS | 1342 | 103 | 5.610E-04 |
| GO_RIBONUCLEOPROTEIN_COMPLEX | 1361 | 97 | 7.800E-03 |

**Table S6. Gene-set analysis for GWAS catalog reported gene sets**

| GeneSet | N | n | adjustEd P |
| --- | --- | --- | --- |
| Body mass index | 1365 | 147 | 1.450E-13 |
| Inflammatory bowel disease | 730 | 89 | 1.160E-10 |
| Heel bone mineral density | 834 | 97 | 1.160E-10 |
| Crohn's disease | 630 | 77 | 2.450E-09 |
| Platelet count | 348 | 51 | 1.180E-08 |
| Bipolar disorder | 656 | 77 | 1.180E-08 |
| Pulse pressure | 690 | 76 | 2.560E-07 |
| Coronary artery disease | 460 | 57 | 3.700E-07 |
| Schizophrenia | 827 | 85 | 5.330E-07 |
| Estimated glomerular filtration rate | 534 | 59 | 7.360E-06 |
| Systolic blood pressure | 793 | 76 | 2.710E-05 |
| Obesity-related traits | 756 | 73 | 3.320E-05 |
| Chronotype | 556 | 58 | 4.180E-05 |
| Ulcerative colitis | 465 | 50 | 9.510E-05 |
| Height | 898 | 80 | 1.950E-04 |
| Diastolic blood pressure | 650 | 57 | 4.880E-03 |
| Blood protein levels | 1935 | 136 | 9.350E-03 |
